# Underdiagnosed Viral Infections in Heart Failure: Metagenomic Sequencing Reveals Association with Left Ventricular Dysfunction

**DOI:** 10.1101/2025.11.11.25339799

**Authors:** Christian Baumeier, Britta Altmann, Gordon Wiegleb, Dominik Harms, Ganna Aleshcheva, C.-Thomas Bock, Sandra Niendorf, Felicitas Escher, Heinz-Peter Schultheiss

**Affiliations:** Institute of Cardiac Diagnostics and Therapy, IKDT GmbH, Berlin, Germany; Robert Koch Institute, Unit 15: Viral Gastroenteritis and Hepatitis Pathogens and Enteroviruses, Department of Infectious Diseases, Berlin, Germany; Institute of Tropical Medicine, University of Tuebingen, Tuebingen, Germany; Deutsches Herzzentrum der Charité, Department of Cardiology, Angiology and Intensive Care Medicine, Berlin, Germany; DZHK (German Centre for Cardiovascular Research), partner site Berlin, Germany

**Author notes:** Corresponding author: Dr. Christian Baumeier, Institute of Cardiac Diagnostics and Therapy, IKDT GmbH, Moltkestrasse 31, 12203, Berlin, Germany.

**Keywords:** Diagnostics, Endomyocardial Biopsy, Heart Failure, Inflammatory Cardiomyopathy, Myocarditis, Pathogen, Virus Detection

## Abstract

**BACKGROUND:** Viral infections are a major cause of inflammatory heart disease, but conventional polymerase chain reaction (PCR) often fails to detect the causative pathogens, limiting diagnostic and therapeutic decisions. We investigated whether targeted metagenomic next-generation sequencing (NGS) improves virus detection in endomyocardial biopsies (EMB) compared to standard PCR, and examined the clinical implications of undetected viral infections in heart failure.

**METHODS:** EMB samples from 108 patients with unexplained heart failure underwent histologic and immunohistochemical analysis to assess myocardial inflammation. All samples were tested for common cardiotropic viruses using PCR. Targeted metagenomic NGS was performed in 36 PCR-positive and 72 PCR-negative cases. Virus prevalence and transcriptional activity were compared with PCR results and correlated with myocardial inflammation and left ventricular ejection fraction (LVEF).

**RESULTS:** NGS identified all 45 viruses previously detected by PCR and revealed 31 additional viral genomes in PCR-positive patients, increasing diagnostic yield by 69%. In PCR-negative patients, NGS identified viral genomes in 56% of cases, uncovering a significant number of previously undiagnosed infections. Frequently detected viruses included parvovirus B19, Epstein-Barr virus, human herpesvirus 6, cytomegalovirus, and adenovirus, as well as less expected agents such as herpesvirus 7/8, adenovirus-associated virus, and pegivirus C. Transcriptionally active parvovirus B19 was more often detected by NGS than PCR (31% vs. 14%). Patients with NGS-confirmed viral infections showed significantly reduced LVEF compared to virus-negative individuals.

**CONCLUSIONS:** Targeted metagenomic sequencing substantially improves virus detection in EMB samples and reveals clinically relevant infections missed by PCR. These results support the integration of NGS into diagnostic workflows for virus-associated heart failure in order to better guide clinical management.

**WHAT IS NEW?:**

- Targeted metagenomic sequencing of endomyocardial biopsies enables comprehensive, broad-range detection of cardiotropic viruses in patients with heart failure.
- Metagenomic sequencing demonstrates markedly higher sensitivity than conventional PCR and can distinguish latent from active viral infections.
- Viral genomes were frequently detected in patients with reduced left ventricular function, indicating a substantial contribution of unrecognized infections to the pathogenesis of heart failure.

**WHAT ARE THE CLINICAL IMPLICATIONS?:**

- Targeted metagenomic sequencing provides a novel diagnostic tool for more accurate etiological classification of heart failure, thereby supporting more informed clinical decision-making.
- Identification of specific viral genomes, including the ability to distinguish latent from active infections, provides a basis for antiviral or immunomodulatory therapy decisions.
- These findings support incorporating metagenomic sequencing into diagnostic workflows to improve risk stratification and management of patients with heart failure.

---

Inflammatory heart disease, encompassing myocarditis and inflammatory cardiomyopathy, is a significant cause of acute and chronic heart failure, particularly among young and middle-aged individuals.^1^ Viral infections are recognized as the leading trigger of myocardial inflammation, with enteroviruses, parvovirus B19, human herpesviruses, and adenoviruses among the most commonly implicated pathogens.^2^ Definitive diagnosis of viral myocardial infection is currently only possible via endomyocardial biopsy (EMB)^3^ and is crucial for guiding clinical management, as the presence or absence of cardiotropic viruses informs different treatment strategies such as antiviral or immunosuppressive therapies.^4,5^

For this purpose, polymerase chain reaction (PCR)-based assays remain the standard diagnostic approach for detecting viral genomes in EMB.^6^ While specific and rapid, conventional PCR is limited by its dependence on a priori target selection, restricting detection to a narrow panel of known pathogens.^7^ Consequently, the true spectrum and prevalence of viral infections in inflammatory heart disease may be substantially underestimated, and cases with false negative PCR results may remain diagnostically unresolved and possibly undergo incorrect treatment.

Advances in metagenomic next-generation sequencing (NGS) offer less biased alternatives to PCR, capable of identifying a broader spectrum of viral genomes, including previously unrecognized or unexpected agents, within clinical tissue samples.^8^ In addition, it offers the possibility to distinguish between latent and active viral infections, which is crucial for treatment decisions.^9^ The utility of NGS in cardiac diagnostics, however, remains underexplored, and its performance relative to standard PCR-based techniques has not been comprehensively evaluated in large patient cohorts with inflammatory heart disease.

Here, we developed a NGS platform for virus detection in EMB and conducted a comparative study to assess the diagnostic yield of NGS versus conventional PCR in patients with heart failure. Our objective was to determine whether NGS improves detection rates of viral genomes and to evaluate the clinical implications of its use in routine diagnostic workflows.

## METHODS

### Study Population

All study participants gave written informed consent, and the study was approved by the Ethic Committee of the Campus Benjamin Franklin (Charité Universitätsmedizin Berlin, EA4/236/20). We enrolled 108 patients presenting with unexplained heart failure that underwent EMB for routine cardiac diagnostics in this study. The cohort comprised 70 male (65%) and 38 female (35%) patients with a mean age of 49 years (interquartile range, 40 to 62). All patients underwent coronary angiography to exclude coronary artery disease and other identifiable causes of myocardial dysfunction. Left ventricular ejection fraction (LVEF) was assessed using the biplane Simpson method by echocardiography.

### Endomyocardial Biopsy and Histologic Evaluation

Six to eight EMB specimens were obtained per patient per procedure. Histologic evaluation was carried out on formalin-fixed, paraffin-embedded tissue using standard staining protocols, including hematoxylin and eosin, Azan, Van Gieson’s, and periodic acid–Schiff stains.^10^ Active myocarditis was diagnosed according to the Dallas criteria and such cases were excluded from the analysis.^11^

### Immunohistochemical Assessment of Myocardial Inflammation

Immunohistochemical analysis was performed on RNAlater-fixed (Thermo Fisher Scientific), cryo-embedded EMB specimens. Myocardial inflammation was defined in accordance with the European Society of Cardiology criteria as the presence of ≥14 CD3+ T lymphocytes per square millimeter.^6^ Additional criteria included thresholds of ≥14 LFA-1+ lymphocytes, ≥40 MAC-1+ macrophages, or ≥40 CD45R0+ memory T cells per square millimeter. Immune cell infiltration was quantified using digital image analysis, as previously described.^10^ Of the 108 patients, 96 (89%) met immunohistochemical criteria for myocardial inflammation, while 12 (11%) did not (Tab. 1).

**Tab. 1:** Patients characteristics and EMB diagnostic results from the analyzed cohort of 108 patients with unclear heart failure.

| **Patients characteristics** | **PCR-positive patients** |  | **PCR-negative patients** |
| --- | --- | --- | --- |
| EMB based diagnosis | DCM | DCMi | DCMi |
| Number, n (%) | 12 (11) | 24 (22) | 72 (67) |
| Age, years (IQR) | 52 (48-59) | 44 (32-58) | 50 (41-62) |
| Sex, n (%) | 6 (50) male<br>6 (50) female | 17 (70) male<br>7 (30) female | 47 (65) male<br>25 (35) female |
| LVEF (%) | 25 ± 13 | 24 ± 14 | 29 ± 12 |
| **EMB inflammation diagnostics** |  |  |  |
| CD3, mean ± SD, cells/mm<sup>2</sup> | 5 ± 5 | 27 ± 43* | 30 ± 20* |
| CD4, mean ± SD, cells/mm<sup>2</sup> | nd | nd | 19 ± 15 |
| CD8, mean ± SD, cells/mm<sup>2</sup> | nd | nd | 12 ± 11 |
| CD45RO, mean ± SD, cells/mm<sup>2</sup> | 23 ± 12 | 82 ± 77* | 76 ± 32* |
| LFA-1, mean ± SD, cells/mm<sup>2</sup> | 6 ± 4 | 39 ± 46* | 41 ± 32* |
| MAC-1, mean ± SD, cells/mm<sup>2</sup> | 13 ± 8 | 38 ± 34* | 58 ± 32* |
| **EMB virus diagnostics** |  |  |  |
| Parvovirus B19, n (%) | 10 (83) | 15 (63) | 0 (0) |
| Latent parvovirus B19, n (%) | 7 (58) | 13 (54) | 0 (0) |
| Active parvovirus B19<sup>#</sup>, n (%) | 3 (25) | 2 (8) | 0 (0) |
| Epstein-Barr virus, n (%) | 3 (25) | 6 (25) | 0 (0) |
| Human herpesvirus 6, n (%) | 4 (33) | 3 (13) | 0 (0) |
| Human cytomegalovirus, n (%) | 1 (8) | 2 (8) | 0 (0) |
| Coxsackievirus B3, n (%) | 0 (0) | 1 (4) | 0 (0) |

\*P<0.05 versus DCM patients; <sup>#</sup>active infection was confirmed by the detection of parvovirus B19 specific viral mRNAs; nd, not determined; DCM, dilated cardiomyopathy; DCMi, inflammatory cardiomyopathy

### Detection of Viral Genomes by Quantitative Polymerase Chain Reaction

Immediately following biopsy, specimens were stabilized in RNAlater (Thermo Fisher Scientific). Genomic DNA was extracted using the Gentra Puregene kit (Qiagen), and total RNA was isolated using TRIzol (Thermo Fisher Scientific) with subsequent DNase treatment (Promega) and complementary cDNA synthesis using random hexamers and the High-Capacity cDNA Reverse Transcription Kit (Thermo Fisher Scientific).^12^ Quantitative polymerase chain reaction (PCR) was used to detect viral genomes, including parvovirus B19 (B19V), enteroviruses (including coxsackievirus B3 (CVB3) and echoviruses), adenoviruses (ADV), Epstein-Barr virus (EBV), human cytomegalovirus (HCMV), and human herpesvirus 6 (HHV-6).^10^ B19V transcriptional activity was assessed via PCR targeting the NS1 and VP1/2 regions.^10,12^

### Targeted Metagenomic Next-Generation Sequencing

Metagenomic analysis was performed on both DNA and RNA fractions obtained from the same extraction procedure as the PCR samples. The RNA was converted into double-stranded cDNA using a previously described two-step synthesis method.^9^ Sequencing libraries were prepared from DNA and cDNA fractions using the NEBNext Ultra II FS DNA Library Prep Kit (New England Biolabs), and enrichment of target sequences for viral species was performed using a custom myBaits^®^ hybridization capture kit according to the manufacturer’s instructions (Daicel Arbor Biosciences). Capture oligonucleotides (length=80bp) were designed using the CATCH tool, optimized for the enrichment of viral genomic and transcriptomic sequences in complex metagenomic samples.^13^ The final bait set comprised 39,574 probe sequences targeting 86 human pathogenic viral species with known or suspected cardiotropism (Suppl. Tab. 1). Following the enrichment of viral DNA or RNA fragments, sequencing was carried out on an Illumina MiSeq^TM^ system with 150 bp paired-end reads (5×10^5^ reads/sample, 300 cycles, v2 reagent kit). Sequence data were processed using Trimmomatic (v0.39), classified taxonomically with Kraken2^14^ (threshold: ≥10 reads), and aligned to reference viral genomes (Suppl. Tab. 2) using bwa-mem2^15^ for DNA reads and HiSat2^16^ for cDNA reads with deduplication and genome coverage analysis via Samtools. Genome coverage visualization was performed using the R package karyoploteR.^17^

### Statistical Analysis

Data are presented as mean ± standard deviation (SD) or median ± SD, unless otherwise indicated. Between group comparisons were conducted using unpaired t-tests. A two-sided exact Fisher’s test was used to test whether the proportions in inflammation-negative and inflammation-positive patients are significantly different. A two-sided P value of ≤ 0.05 was considered to indicate statistical significance. All analyses were conducted using GraphPad Prism, version 10.3.1 (GraphPad Software).

## RESULTS

### Patients Characteristics and Biopsy Findings

EMB from a total of 108 patients with unexplained heart failure were analyzed for viral infections using both conventional PCR-based diagnostics and the novel targeted metagenomic NGS tool. Two thirds of the patients (n=70, 65%) were male and the mean age was 49±16 years. The mean LVEF was 28±12%, and in 75% (n=80) of cases the LVEF was reduced to ≤ 45%. The total cohort was divided into viral PCR-positive (n=36) and PCR-negative (n=72) patients according to the results of the routine PCR-based diagnostic setting (Tab. 1). In 24 of the PCR-positive patients and in all of the PCR-negative patients, histological and immunohistochemical diagnostics revealed evidence of increased infiltration of immune cell, leading to the diagnosis of inflammatory cardiomyopathy (Fig. 1, Tab. 1). As acute myocarditis cases were excluded from this study, enrolled patients did not present with cardiomyocyte necrosis.

**Figure 1:**
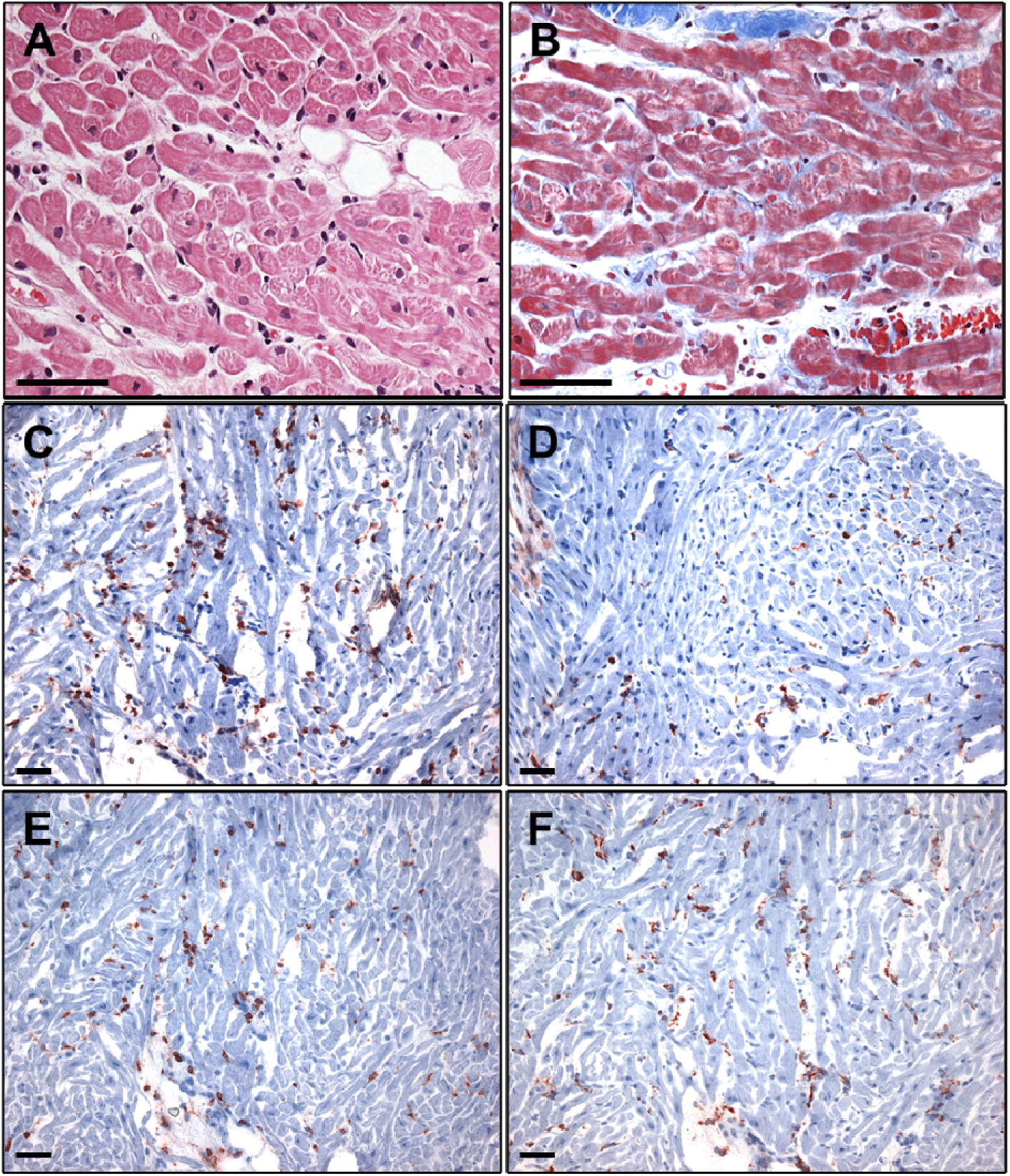
Representative histological and immunohistochemical images of inflammatory cardiomyopathy. **(A)** Hematoxylin and eosin and **(B)** Azan histological staining. **(C-D)** Immunohistochemical staining shows increased infiltration of CD3+ T cells (C), CD45R0+ memory T cells (D), LFA-1+ lymphocytes (E), and MAC-1+ macrophages (F). Magnification: 400x (A, B), 200x (C-F). Scale bar = 50 µm.

In 12 of the PCR-positive patients, no signs of inflammation were found, but enlargement of the left ventricle was observed, leading to the diagnosis of dilated cardiomyopathy (DCM) (Tab. 1). All EMB showed mild to moderate fibrosis and normotrophic up to mildly hypertrophic cardiomyocytes (Fig. 1). Among the viruses detected by PCR were parvovirus B19 (B19V), Epstein-Barr virus (EBV), human herpesvirus 6 (HHV-6), human cytomegalovirus (HCMV) and coxsackievirus B3 (CVB3) (Fig. 2, Tab. 1). In B19V-positive cases, the infection was classified as latent or active based on the detection of B19V-specific RNA transcripts (Tab. 1).

**Figure 2:**
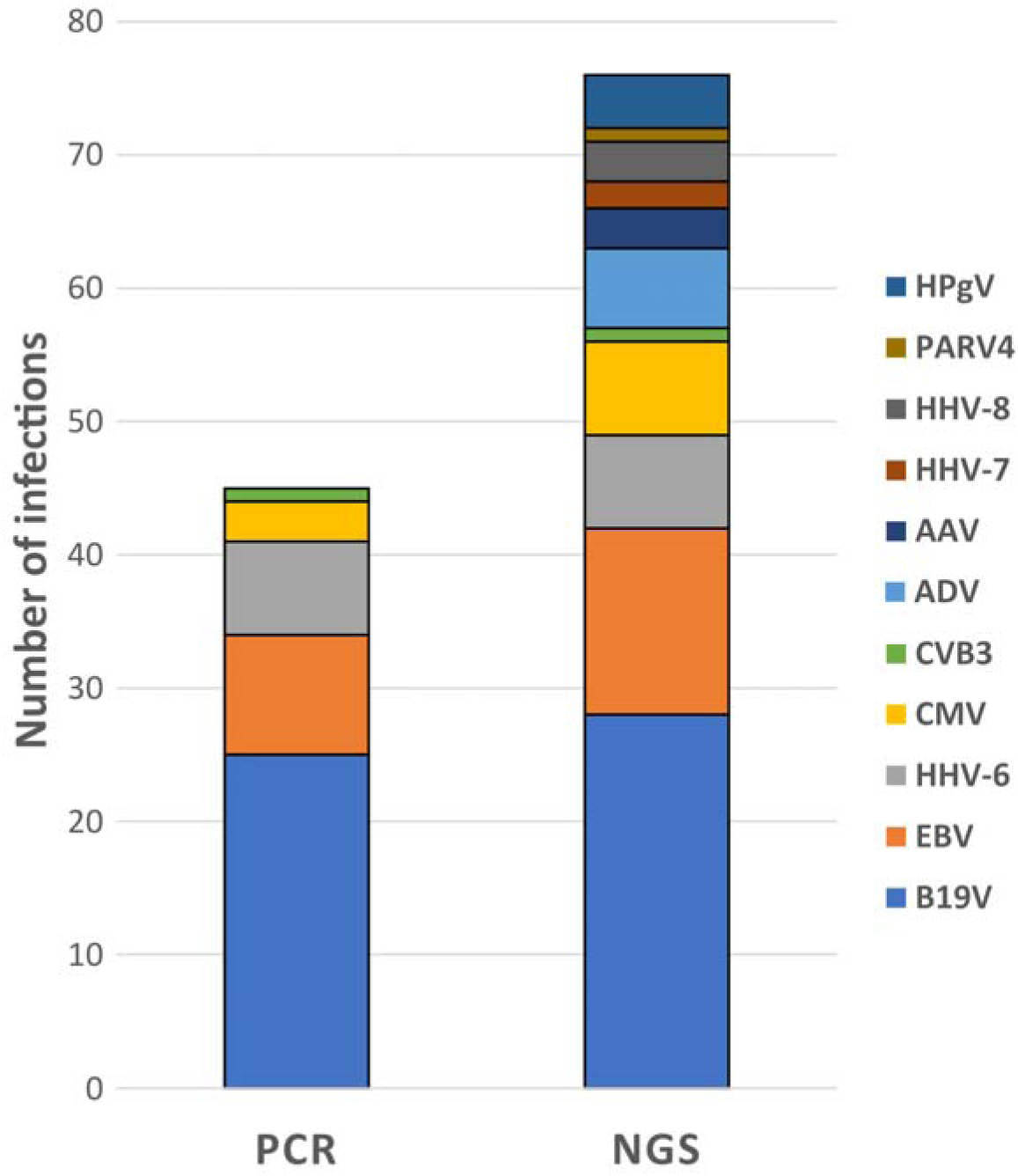
Number of detected viral genomes by PCR and NGS in the same EMB samples from 36 PCR-positive patients. B19V, parvovirus B19; EBV, Epstein-Barr virus; HHV-6, human herpesvirus 6; HCMV, human cytomegalovirus; ADV, adenovirus; CVB3, coxsackievirus B3; AAV, adeno-associated virus; HHV-7, human herpesviruses 7; HHV-8, human herpesviruses 8; PARV4, tetraparvovirus; HPgV, pegivirus C.

### Targeted Metagenomic Next-Generation Sequencing Demonstrates Increased Diagnostic Performance Compared to PCR Diagnostics

To determine the diagnostic performance of the NGS tool, a cohort of 36 patients with previously PCR-confirmed infections was analyzed by NGS. The viruses detected using the conventional PCR method were B19V (n=25; 69%), EBV (n=9; 25%), HHV-6 (n=7; 19%), HCMV (n=3; 8%) and CVB3 (n=1; 3%) (Tab. 2). An active B19V infection was confirmed in five (14%) of the cases by the detection of viral VP1/2- or NS1-specific mRNA (Tab. 2).

**Tab. 2:** Number of viral genomes detected in 36 EMB by PCR and NGS diagnostics.

| Species | PCR Diagnostics | NGS Diagnostics | Additional Infections by NGS |
| --- | --- | --- | --- |
| Parvovirus B19, n (%) | 25 (69) | 28 (78) | 3 (9) |
| Latent parvovirus B19, n (%) | 20 (56) | 17 (47) | 0 (0) |
| Active parvovirus B19<sup>#</sup>, n (%) | 5 (14) | 11 (31) | 6 (17) |
| Epstein-Barr virus, n (%) | 9 (25) | 14 (39) | 5 (14) |
| Human herpesvirus 6, n (%) | 7 (19) | 7 (19) | 0 (0) |
| Human cytomegalovirus, n (%) | 3 (8) | 7 (19) | 4 (11) |
| Adenovirus, n (%) | 0 (0) | 6 (17) | 6 (17) |
| Coxsackievirus B3, n (%) | 1 (3) | 1 (3) | 0 (0) |
| Adeno-Associated Virus, n (%) | - | 3 (8) | 3 (8) |
| Human herpesvirus 7, n (%) | - | 2 (6) | 2 (6) |
| Human herpesvirus 8, n (%) | - | 3 (8) | 3 (8) |
| Tetraparvovirus, n (%) | - | 1 (3) | 1 (3) |
| Pegivirus C, n (%) | - | 4 (11) | 4 (11) |
| **Total number of infections, n** | **45** | **76** | **31** |

<sup>#</sup> not routinely measured by PCR; <sup>#</sup>active infection was confirmed by the detection of parvovirus B19 specific mRNA

The analysis of the same EMB samples with our NGS platform confirmed all previously identified viral genomes (n=45) and could detect 31 additional infections, representing a 69% increase in diagnostic yield (Fig. 2, Tab. 2). The viral genomes identified by the NGS approach included a higher number of known cardiotropic pathogens such as B19V (n=28; 78%), EBV (n=14; 39%), HHV-6 (n=7; 19%), HCMV (n=7; 19%), adenovirus (ADV; n=6; 17%) and CVB3 (n=1; 3%), indicating a higher incidence of these pathogens than previously diagnosed by conventional methods (Fig. 2, Tab. 2). Importantly, the number of active B19V cases was also increased in the NGS analysis (31% vs. 14%), resulting in a higher incidence of B19V transcriptional activity than assumed (Tab. 2). In patients previously diagnosed as latent, viral RNA was retrospectively detected using NGS, leading to a shift toward transcriptionally active B19V infections (Tab. 2). In addition to the known cardiotropic viruses, genomes of adeno-associated virus (AAV; n=3; 8%), human herpesvirus 7 (HHV-7; n=2; 6%), human herpesvirus 8 (HHV-8; n=3; 8%), tetraparvovirus (PARV4; n=1; 3%), and pegivirus C (HPgV; n=4; 11%) were detected by NGS (Fig. 2, Tab. 2). These viruses have not been examined in the setting of routine PCR-based diagnostics.

Interestingly, infections with viruses such as ADV, HHV-8, and PARV4 were found exclusively in patients with myocardial inflammation, with significant differences for ADV and HHV-8 (Suppl. Fig. 1). In comparison, other viruses were evenly distributed between patients with and without inflammation, with EBV occurring significantly more frequently in patients without inflammation (Suppl. Fig. 1). An additional comparison between patients with and without inflammation showed that although both groups had the same average number of infections, patients with inflammation were more likely to have multiple infections (Suppl. Fig. 2). Thus, a total of 10 patients with multiple infections (>3) were identified using NGS, while a maximum of three co-infections could be detected in a sample using PCR.

### Targeted Metagenomic Next-Generation Sequencing Reveal a High Proportion of Previously Undiagnosed Viral Infections

The discovery of additional infections in the PCR-positive cohort has led to the hypothesis that a significant number of patients with previously determined virus-negative EMB and myocardial inflammation could actually test positive for the presence of viral genomes or transcripts within the myocardium using the new metagenomic NGS methodology. Therefore, we next subjected a cohort of 72 patients with inflammatory cardiomyopathy and negative viral PCR results to the NGS analysis. In 31 (44%) patients, no virus could be detected with NGS, while 40 (56%) of the tested EMBs were positive in the NGS analysis (Fig. 3, Tab. 3). A comparison of the age and gender distribution between NGS-positive and NGS-negative patients revealed no differences, however NGS-positive patients had a significantly lower LVEF (Tab. 3). The inflammation parameters did not differ between these groups and were elevated in all patients (Tab. 3). Among the 56 additional viral genomes detected by NGS were known cardiotropic pathogens such as B19V, HHV-6, EBV, ADV, and HCMV, but also viruses that are not routinely tested for, such as AAV, HHV-8, HPgV, PARV4, and herpes simplex virus 1 (HSV-1) (Fig. 3, Tab. 3).

**Figure 3:**
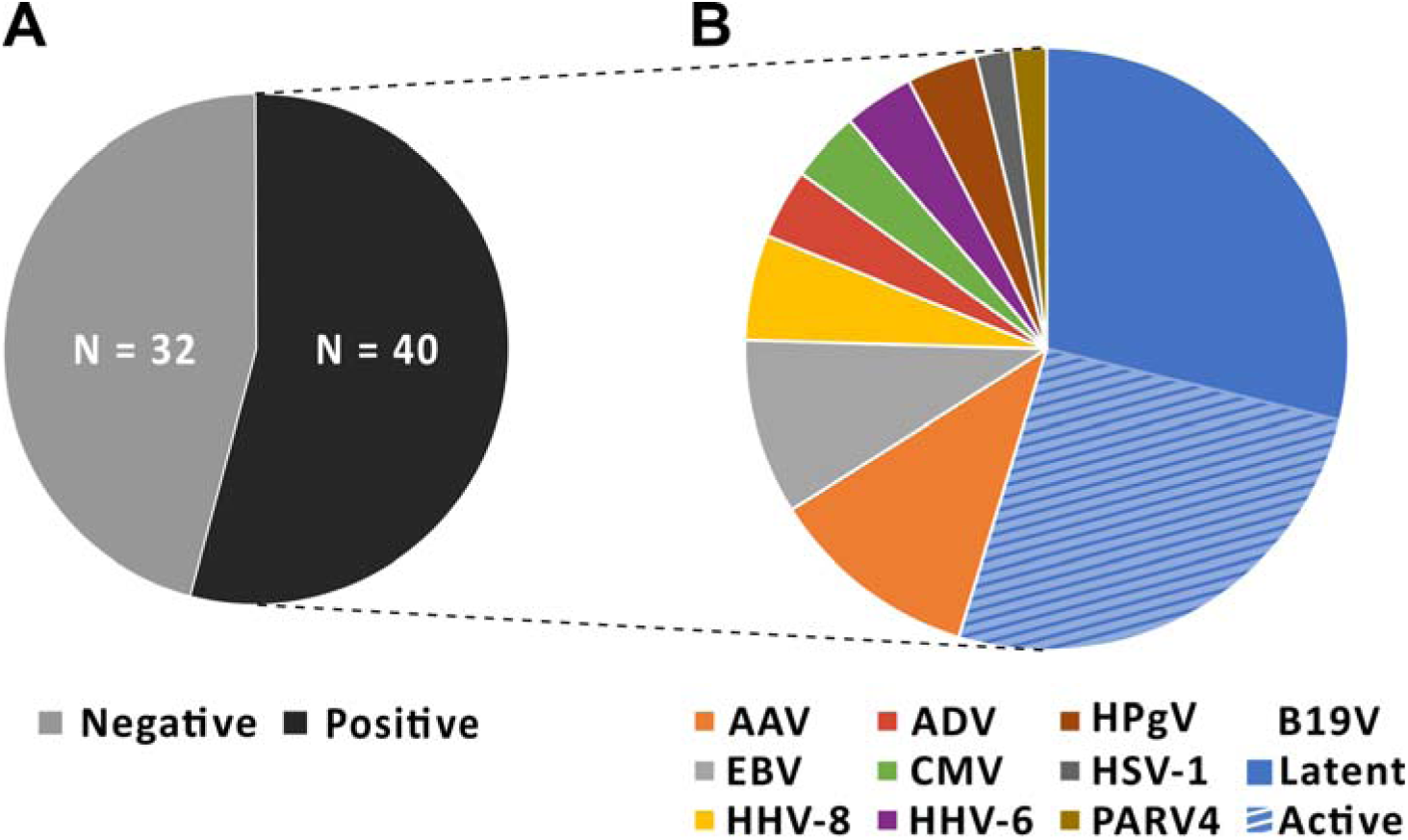
Results from targeted metagenomic NGS analysis of 72 patients with inflammatory cardiomyopathy who had previously tested negative for the presence of viral genomes or transcripts using PCR. **(A)** Viral genomes/transcripts were detected using NGS in 40 (56%) of 72 tested PCR-negative cases. **(B)** Proportion of viral genomes/transcripts detected in the 40 NGS-positive EMB. Differentiation of active and latent B19V infections was performed using RNA transcript analysis. B19V, parvovirus B19; EBV, Epstein-Barr virus; HHV-6, human herpesvirus 6; HCMV, human cytomegalovirus; ADV, adenovirus; CVB3, coxsackievirus B3; AAV, adeno-associated virus; HSV-1, herpes simplex virus 1; HHV-8, human herpesviruses 8; PARV4, tetraparvovirus; HPgV, pegivirus C.

**Tab. 3:** Patients characteristics and EMB results from the analyzed cohort of 72 patients with in-flammatory cardiomyopathy and negative PCR results.

| Patients' characteristics | All | NGS-negative | NGS-positive | P-value |
| --- | --- | --- | --- | --- |
| Number, n (%) | 72 (100) | 32 (44) | 40 (56) |  |
| Age, years (IQR) | 50 (41-62) | 56 (40-62) | 48 (42-62) | 0.526 |
| Sex, n (%) | 47 (65) male<br>25 (35) female | 22 (68) male<br>10 (32) female | 25 (62) male<br>15 (38) female | 0.638 |
| LVEF (%) | 29 ± 12 | 32 ± 12 | 26 ± 10 | **0.039** |
| **EMB inflammation diagnostics** |  |  |  |  |
| CD3, mean ± SD, cells/mm<sup>2</sup> | 30 ± 20 | 32 ± 20 | 29 ± 20 | 0.552 |
| CD4, mean ± SD, cells/mm<sup>2</sup> | 19 ± 15 | 23 ± 17 | 14 ± 11 | 0.103 |
| CD8, mean ± SD, cells/mm<sup>2</sup> | 12 ± 11 | 14 ± 11 | 9 ± 11 | 0.273 |
| CD45R0, mean ± SD, cells/mm<sup>2</sup> | 76 ± 32 | 78 ± 28 | 78 ± 39 | 0.988 |
| LFA-1, mean ± SD, cells/mm<sup>2</sup> | 41 ± 32 | 39 ± 32 | 42 ± 33 | 0.675 |
| MAC-1, mean ± SD, cells/mm<sup>2</sup> | 58 ± 32 | 66 ± 33 | 52 ± 30 | 0.086 |
| **EMB virus diagnostics** |  |  |  |  |
| Total infections, n | 56 |  | 56 | - |
| Parvovirus B19, n (%) | 32 (44) |  | 32 (44) | - |
| Latent parvovirus B19, n (%) | 17 (24) |  | 17 (24) | - |
| Active parvovirus B19<sup>#</sup>, n (%) | 15 (21) |  | 15 (21) | - |
| Adeno-Associated Virus, n (%) | 6 (8) |  | 6 (8) | - |
| Epstein-Barr virus, n (%) | 5 (7) |  | 5 (7) | - |
| Human herpesvirus 8, n (%) | 3 (4) | 0 (0) | 3 (4) | - |
| Human herpesvirus 6, n (%) | 2 (3) |  | 2 (3) | - |
| Human cytomegalovirus, n (%) | 2 (3) |  | 2 (3) | - |
| Adenovirus, n (%) | 2 (3) |  | 2 (3) | - |
| Pegivirus C, n (%) | 2 (3) |  | 2 (3) | - |
| Herpes simplex virus 1, n (%) | 1 (1) |  | 1 (1) | - |
| Tetraparvovirus, n (%) | 1 (1) |  | 1 (1) | - |

<sup>#</sup>active infection was confirmed by the detection of parvovirus B19 specific mRNA

### Targeted Metagenomic Next-Generation Sequencing Enables the Genotypic Characterization of Viral Pathogens

In EMB from nine patients, we obtained nearly complete viral genome coverage (>80%), which provided us with important information about the genotype and origin of the detected strain. In a female patient with elevated lymphocytic inflammation, no infection was detected by PCR, but the NGS approach confirmed a high viral load of B19V. The sequences yielded genome coverage of >99% (4985 bp, Fig. 4A, Suppl. Tab. 2), and phylogenetic analysis revealed a match with erythrovirus V9 (B19V genotype 3), a B19V variant that has rarely been associated with inflammatory heart disease to date.^18^ In addition, NGS was able to detect the complete genome sequence (<99% genome coverage, 6882 bp) of an enterovirus B strain in a patient who died shortly after EMB (Fig. 4B, Suppl. Tab. 2). For more complex viral genomes such as EBV, sequence alignment achieved up to 61% genome coverage (104.8 Kbp, Fig. 4C, Suppl. Tab. 2).

**Figure 4:**
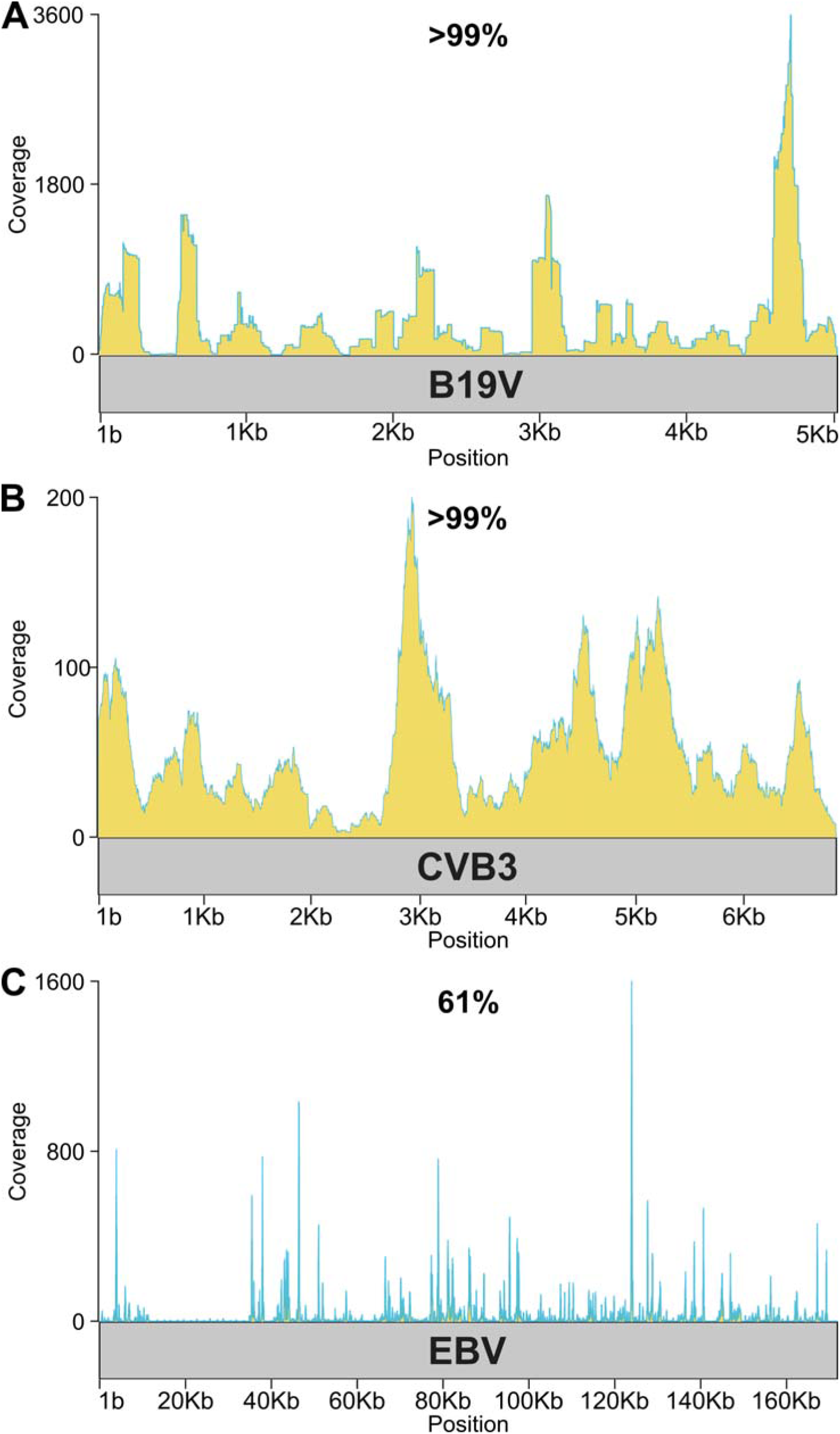
Genome coverage plots showing read depth of patients with **(A)** parvovirus B19 (B19V, >99% of the genomic sequence covered), **(B)** coxsackievirus B3 (CVB3, >99% of the genomic sequence covered), and **(C)** Epstein-Barr virus (EBV, 61% of the genomic sequence covered).

## DISCUSSION

In this study, we evaluated the diagnostic performance of a novel NGS platform using EMB from patients with unexplained heart failure, comparing it directly with paired data from conventional PCR-based viral diagnostics. Our findings demonstrate a markedly superior performance of the NGS methodology in detecting viral genomes, including both known cardiotropic pathogens and previously unrecognized or untested viral species. This increased capacity not only led to the identification of a broader viral spectrum in the myocardium but also revealed a considerable number of viral infections in patients previously classified as virus-negative using standard PCR-based diagnostics in EMB. This has significant implications for treatment decisions and could revolutionize the molecular diagnostics of inflammatory heart disease.

### Targeted Metagenomic Next-Generation Sequencing Improves the Detection of Viral Infections in the Myocardium

Among patients with previously PCR-confirmed infections, NGS detected all viruses identified by PCR and uncovered an additional 31 infections (69% increase), expanding the viral landscape to include adeno-associated virus (AAV), human herpesviruses 7 and 8 (HHV-7, HHV-8), tetraparvovirus (PARV4), and pegivirus C (HPgV), which are not routinely assessed. Importantly, NGS analysis detects a higher prevalence of active B19V infections than PCR, suggesting a greater incidence of B19V transcription than previously assumed.^2^ Using the novel NGS approach, both viral DNA and RNA are enriched and sequenced across the entire B19V genome, resulting in greater detection of viral transcripts and thus a higher probability of detecting active infections. The prognostic and therapeutic significance of transcriptionally active B19V infection has already been demonstrated in a cohort of 871 consecutive B19V-positive patients with non-ischemic cardiomyopathy.^10^ The data presented here suggest a large number of undetected active B19V infections, which indicates the need of specific antiviral therapy over generalized immunosuppression.^19,20^

Previously undetected viruses such as ADV and HHV-8 were identified exclusively in patients with myocardial inflammation, indicating a potential association with the inflammatory process in myocardial disease. Since persistent ADV is associated with progressive deterioration of LVEF, the detection and treatment of ADV infection are crucial for the clinical outcome of patients.^21,22^ Interestingly, EBV genomes were found more often in non-inflamed myocardium, consistent with its known ability to establish latent infections with variable pathogenic relevance.^23^ In this context, we have already been able to show that the NGS approach can be used to classify EBV infections as lytic or latent, which has an impact on the extent of inflammation.^9^ Our RNA sequencing data indicate a slightly higher but not significant prevalence of lytic EBV transcripts in patients with inflammation, which needs to be investigated in a larger EBV-positive cohort.

### Targeted Metagenomic Next-Generation Sequencing Identifies Numerous Undetected Viral Infections

Among PCR-negative patients with biopsy-proven inflammatory cardiomyopathy, NGS identified viral genomes in over half the cases (56%). This finding has significant clinical and therapeutic implications, as these patients would otherwise be misclassified as virus-negative and potentially managed without consideration of a viral etiology. Interestingly, NGS-positive patients exhibited significantly lower LVEF compared to NGS-negative counterparts, which may indicate more advanced or aggressive disease associated with persistent viral infection.^21^ High-resolution virus diagnostics are particularly important in order to assess which patients can undergo immunosuppressive therapy, as this is contraindicated for virus-positive patients.^4,24^ Such patients were more likely to exhibit multiple infections, highlighting the possible additive or synergistic effects of co-infections in myocardial damage and dysfunction.^5^

Metagenomic analyses in cardiomyopathy patients have been limited to date. Apart from a few case reports focusing on the analysis of blood samples, there are no large-scale studies using NGS in cardiac tissue samples. Takeuchi et al. applied NGS to serum samples from patients with myocarditis and identified viral genetic material in 41% of cases, underscoring the potential usefulness of this method in detecting viral pathogens that may not be detectable using conventional methods.^25^ The identified species included B19V, EBV, HPgV, torque teno virus, and respiratory syncytial virus. Another study by Heidecker et al. investigated viral infection in myocarditis patients using metagenomic NGS of peripheral mononuclear blood cells (n=24), plasma (n=27), EMB (n=2), and explanted hearts (n=13).^26^ In the blood samples, they found genomes of B19V, EBV, HCMV, HPgV, anellovirus, and retrovirus K (HERVK), while only HERVK and small amounts of B19V, EBV, and HCMV were detected in the tissue samples. However, the small number of EMB in this study limits the conclusions about viral persistence in the myocardium. To the best of our knowledge, this is the first study to apply a metagenomic NGS approach to EMB from 108 heart failure patients, providing a first comprehensive mapping of cardiotropic viruses in myocardial disease.

### Diagnostic and Therapeutic Considerations

The improved diagnostics achieved through targeted metagenomic NGS underscores the limitations of standard PCR, which relies on prior selection of short viral genomic or transcriptional targets. In contrast, our NGS approach offers near-complete viral genome coverage in the selected viral species and the ability to uncover unexpected viral agents, enabling insights into strain variation, classification, and potential antiviral resistance.^13^ For example, we identified a B19V strain related to genotype 3 (erythrovirus V9), which may differ in tropism or pathogenicity compared to more common variants.^27^ In addition, nearly complete genome coverage was achieved for a CVB3 infection, leading to the identification of a CVB3 strain that had originally been isolated from clinical stool samples.^28^ Since this patient died shortly after EMB analysis, this strain may be more pathogenic than other enterovirus strains, but this needs to be investigated in further studies.

Importantly, establishing virus-positivity or -negativity within patient myocardium is essential for treatment decisions in myocarditis and inflammatory cardiomyopathy.^1^ Immunosuppressive therapy can be harmful in patients with active viral infection, whereas those with virus-negative EMB benefit substantially from immunosuppression, with improved left ventricular function and long-term outcomes.^4,29,30^ On the other hand, virus-positive patients derive less benefit, and potentially suffer harm from immunosuppression, whereas antiviral therapy may be more appropriate.^1,2,5^ Indeed, the European Society of Cardiology endorses EMB with viral genome testing both to confirm etiology and to guide therapy: immunosuppression only in virus-negative cases, and antiviral or supportive strategies in virus-positive patients with myocardial inflammation.^6,31^ Although large trials of antiviral treatment remain limited, interferon-β and nucleoside analogues have shown some promise for clearing viral genomes and improving cardiac function in chronic viral cardiomyopathy.^19,20,22,32^ The use of antiherpetic agents has been considered for patients with myocardial inflammation associated with HSV-1, HHV-6, EBV, or HCMV, although definitive evidence of their clinical efficacy remains lacking.^33,34^

Our findings support incorporating NGS into the EMB diagnostic workflow to achieve precise pathogen detection, enabling robust etiological classification. This can help distinguish active from latent infections, an essential distinction for choosing between antiviral therapy and immunosuppression. By enabling personalized and pathogen-directed treatment, NGS-enhanced diagnostics may improve clinical outcomes and avoid inappropriate treatment in patients with inflammatory cardiomyopathies.

### Study Limitations

Although NGS demonstrated superior diagnostic performance compared to conventional PCR, the clinical significance of several viruses, such as pegivirus C, tetraparvovirus, and adenovirus-associated virus, remains unclear and requires further investigation to clarify their role in inflammatory heart disease. Furthermore, NGS implementation is currently limited by factors such as cost, longer turnaround times, and the need for specialized bioinformatics infrastructure. Longitudinal studies integrating clinical follow-up and therapeutic outcomes will be crucial to determine the prognostic and therapeutic value of NGS-guided virus diagnostics in inflammatory heart disease.

## COCLUSIONS

The present study shows that NGS-based virus diagnostics significantly improves the detection of known cardiotropic viruses and viruses that are not routinely tested for in EMBs from patients with unexplained heart failure. The method outperforms conventional PCR in terms of virus detection rate, detection of active viral transcription, and breadth of viral spectrum, and identifies additional infections in both previously PCR-positive and PCR-negative patients. Importantly, NGS-confirmed viral infections were associated with significantly reduced left ventricular ejection fraction, underscoring their clinical relevance. These findings support the integration of targeted metagenomic NGS into routine diagnostic workflows for the etiological characterization of inflammatory heart diseases, with the aim of enabling causal, specific, and personalized treatment.

## Data Availability

All data mentioned in the manuscript are available upon request.

## Acknowledgments

We thank Kitty Winter, Susanne Ochmann, Jenny Klostermann, Ivana Zapper, Mona Fechler-Bitteti, and Hannes Jarmuth for their technical assistance.

## Supplemental Material

**Suppl. Fig. 1:**
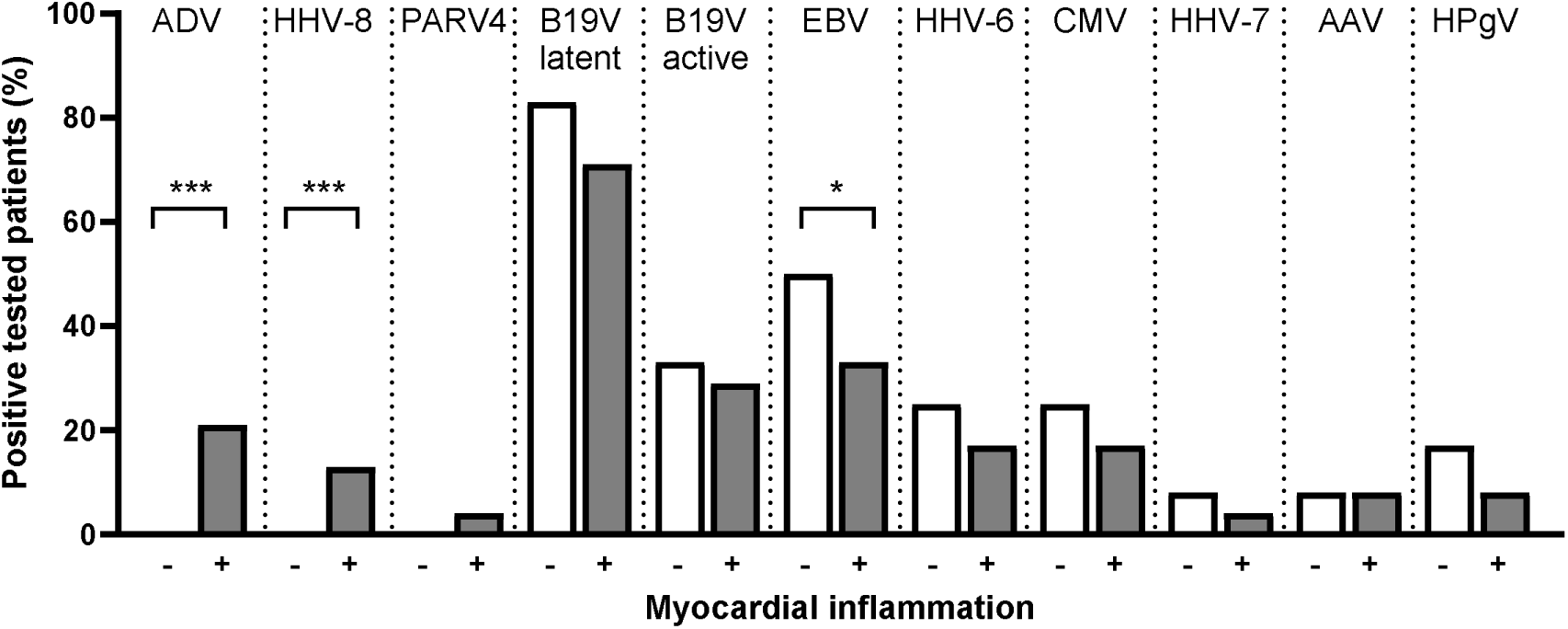
Proportion of viral infections according to NGS analysis in patients without inflammation (−) compared to patients with inflammation (+). A two-sided exact Fisher’s test was used for statistical comparison. *P < 0.05; ***P < 0.001.

**Suppl. Fig. 2:**
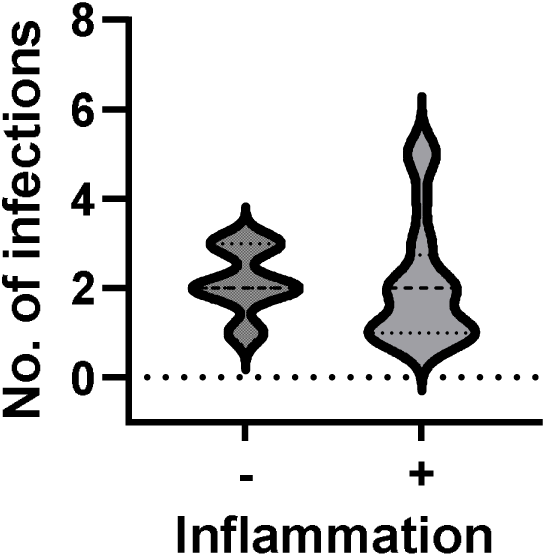
Violin plot showing the total number of viral infections after NGS analysis in patients without inflammation (−) and with inflammation (+).

**Suppl. Tab. 1:**
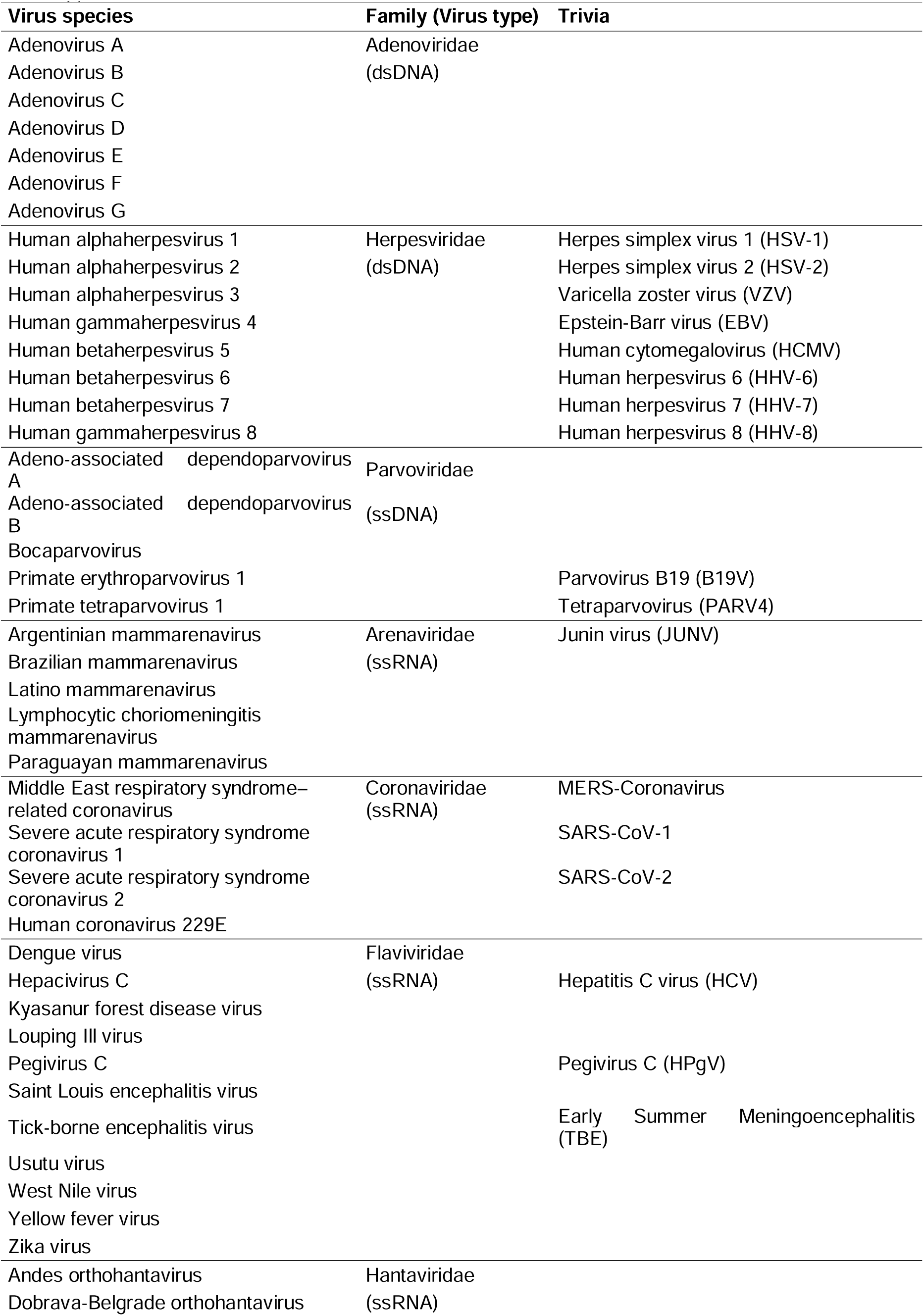
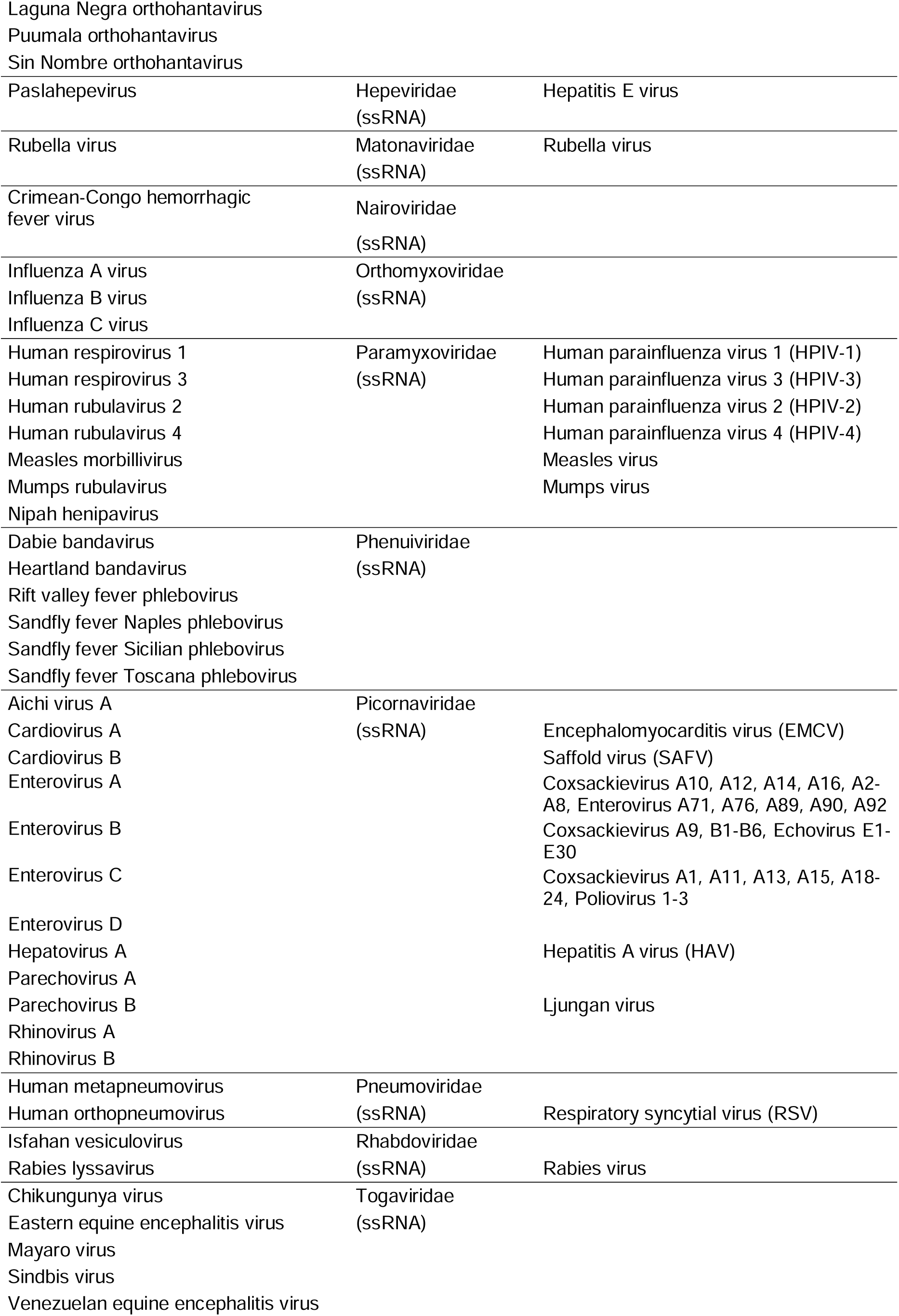
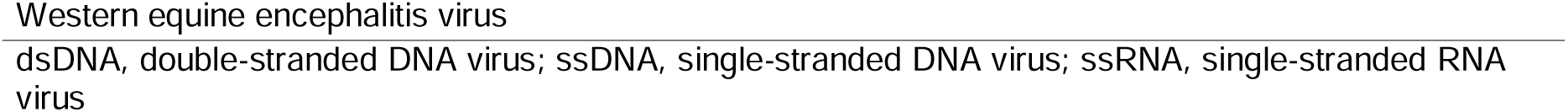
Overview of all 86 virus species that can be enriched and detected using the NGS approach.

**Suppl. Tab. 2:** Reference virus genomes with the corresponding genome size and maximum cover-age of the mNGS data in the alignment.

| Virus species | Reference genomes | Genome size<br>($\times 10^3$ bp) | Maximum genome<br>coverage (%) |
| --- | --- | --- | --- |
| Adeno-associated virus | NC_001401 | 4.68 | 28.0 |
| Adenovirus | NC_003266; NC_001405 | 35.9; 35.9 | 1.9 |
| Parvovirus B19 | AY044266.2; AY386330;<br>NC_000883; NC_004295 | 4.94; 5.59;<br>5.59; 5.03 | >99.0 |
| Cytomegalovirus | NC_006273 | 235.6 | 1.5 |
| Coxsackievirus B3 | MF678304 | 6.9 | >99.0 |
| Epstein-Barr virus | NC_009334; NC_007605 | 172.8; 171.8 | 61.0 |
| Human herpesvirus 6 | NC_001664; NC_000898 | 159.4; 162.1 | 2.5 |
| Human herpesvirus 7 | NC_001716 | 153.1 | 0.6 |
| Human herpesvirus 8 | NC_009333 | 137.9 | 0.5 |
| Pegivirus C | NC_001710 | 9.39 | 13.4 |
| Herpes simplex virus 1 | NC_001806 | 152.2 | 0.7 |
| Tetraparvovirus | NC_007018 | 5.27 | 41.3 |

## References

1. Ammirati E, Frigerio M, Adler ED, Basso C, Birnie DH, Brambatti M, Friedrich MG, Klingel K, Lehtonen J, Moslehi JJ, Pedrotti P, Rimoldi OE, Schultheiss H-P, Tschöpe C, Cooper LT, Camici PG. Management of Acute Myocarditis and Chronic Inflammatory Cardiomyopathy. Circ Hear Fail [Internet]. 2020;13. Available from: https://www.ahajournals.org/doi/10.1161/CIRCHEARTFAILURE.120.007405

2. Baumeier C, Harms D, Aleshcheva G, Gross U, Escher F, Schultheiss H-P. Advancing Precision Medicine in Myocarditis: Current Status and Future Perspectives in Endomyocardial Biopsy-Based Diagnostics and Therapeutic Approaches. J Clin Med [Internet]. 2023;12:5050. Available from: https://www.mdpi.com/2077-0383/12/15/5050

3. Schulz-Menger J, Collini V, Gröschel J, Adler Y, Brucato A, Christian V, Ferreira VM, Gandjbakhch E, Heidecker B, Kerneis M, Klein AL, Klingel K, Lazaros G, Lorusso R, Nesukay EG, Rahimi K, Ristić AD, Rucinski M, Sade LE, Schaubroeck H, Semb AG, Sinagra G, Thune JJ, Imazio M, Arbelo E, Basso C, Adamo M, Aktaa S, Ammirati E, Anderson L, Arbustini E, Bobbio E, Boriani G, Brida M, Byrne RA, Caforio ALP, Dan G-A, Domínguez F, Fredericks S, Gulati G, Ibanez B, James S, Kharlamov A, Klaassen S, Kluin J, Koskinas KC, Kuchynka P, Kunadian V, Landmesser U, Lip GYH, Maisch B, Marelli-Berg F, Martin P, McEvoy JW, Mihaylova B, Mindham R, Moelgaard I, Mohiddin SA, Nielsen JC, Pasquet AA, Peretto G, Pilichou K, Piriou N, Prescott E, Rakisheva A, Rocca B, Rossello X, Sannino A, Seidel F, Tanner FC, Tomkowski WZ, Vaartjes I, Van Linthout S, Vrints C, Wojnicz R, Zeppenfeld K, Banushi AD, Chettibi M, Sisakian HS, Musayev O, Paelinck BP, Begić A, Daskalov Y, Skoric B, Ioannides M, Palecek T, Rossing K, Ghareeb HS, Hinto U, Kupari M, Berthelot E, Agladze V, Gerull B, Kasiakogias A, Vágó H, Ingimarsdóttir IJ, Joyce E, Goland S, et al. 2025 ESC Guidelines for the management of myocarditis and pericarditis. Eur Heart J [Internet]. 2025;Available from: https://academic.oup.com/eurheartj/advance-article/doi/10.1093/eurheartj/ehaf192/8234483

4. Frustaci A, Chimenti C. Immunosuppressive Therapy in Myocarditis. Circ J [Internet]. 2014;79:4–7. Available from: https://www.jstage.jst.go.jp/article/circj/79/1/79_CJ-14-1192/_article

5. Schultheiss H-P, Baumeier C, Aleshcheva G, Bock C-T, Escher F. Viral Myocarditis—From Pathophysiology to Treatment. J Clin Med [Internet]. 2021;10:5240. Available from: https://www.mdpi.com/2077-0383/10/22/5240

6. Caforio ALP, Pankuweit S, Arbustini E, Basso C, Gimeno-Blanes J, Felix SB, Fu M, Heliö T, Heymans S, Jahns R, Klingel K, Linhart A, Maisch B, McKenna W, Mogensen J, Pinto YM, Ristic A, Schultheiss H-P, Seggewiss H, Tavazzi L, Thiene G, Yilmaz A, Charron P, Elliott PM, European Society of Cardiology Working Group on Myocardial and Pericardial Diseases. Current state of knowledge on aetiology, diagnosis, management, and therapy of myocarditis: a position statement of the European Society of Cardiology Working Group on Myocardial and Pericardial Diseases. Eur Heart J [Internet]. 2013;34:2636–48, 2648a-2648d. Available from: http://www.ncbi.nlm.nih.gov/pubmed/23824828

7. Seferović PM, Tsutsui H, McNamara DM, Ristić AD, Basso C, Bozkurt B, Cooper LT, Filippatos G, Ide T, Inomata T, Klingel K, Linhart A, Lyon AR, Mehra MR, Polovina M, Milinković I, Nakamura K, Anker SD, Veljić I, Ohtani T, Okumura T, Thum T, Tschöpe C, Rosano G, Coats AJS, Starling RC. Heart Failure Association of the ESC, Heart Failure Society of America and Japanese Heart Failure Society Position statement on endomyocardial biopsy. Eur J Heart Fail [Internet]. 2021;23:854–871. Available from: https://onlinelibrary.wiley.com/doi/10.1002/ejhf.2190

8. Tang H, Chen Y, Tang X, Wei M, Hu J, Zhang X, Xiang D, Yang Q, Han D. Yield of clinical metagenomics: insights from real-world practice for tissue infections. eBioMedicine [Internet]. 2025;111:105536. Available from: https://linkinghub.elsevier.com/retrieve/pii/S2352396424005723

9. Baumeier C, Harms D, Altmann B, Aleshcheva G, Wiegleb G, Bock T, Escher F, Schultheiss H-P. Epstein-Barr Virus Lytic Transcripts Correlate with the Degree of Myocardial Inflammation in Heart Failure Patients. Int J Mol Sci [Internet]. 2024;25:5845. Available from: https://www.mdpi.com/1422-0067/25/11/5845

10. Escher F, Aleshcheva G, Pietsch H, Baumeier C, Gross UM, Schrage BN, Westermann D, Bock C-T, Schultheiss H-P. Transcriptional Active Parvovirus B19 Infection Predicts Adverse Long-Term Outcome in Patients with Non-Ischemic Cardiomyopathy. Biomedicines [Internet]. 2021;9:1898. Available from: https://www.mdpi.com/2227-9059/9/12/1898

11. Aretz HT. Myocarditis: the Dallas criteria. Hum Pathol [Internet]. 1987;18:619–24. Available from: http://www.ncbi.nlm.nih.gov/pubmed/3297992

12. Pietsch H, Escher F, Aleshcheva G, Lassner D, Bock C-T, Schultheiss H-P. Detection of parvovirus mRNAs as markers for viral activity in endomyocardial biopsy-based diagnosis of patients with unexplained heart failure. Sci Rep [Internet]. 2020;10:22354. Available from: http://www.nature.com/articles/s41598-020-78597-4

13. Metsky HC, Siddle KJ, Gladden-Young A, Qu J, Yang DK, Brehio P, Goldfarb A, Piantadosi A, Wohl S, Carter A, Lin AE, Barnes KG, Tully DC, Corleis B, Hennigan S, Barbosa-Lima G, Vieira YR, Paul LM, Tan AL, Garcia KF, Parham LA, Odia I, Eromon P, Folarin OA, Goba A, Simon-Lorière E, Hensley L, Balmaseda A, Harris E, Kwon DS, Allen TM, Runstadler JA, Smole S, Bozza FA, Souza TML, Isern S, Michael SF, Lorenzana I, Gehrke L, Bosch I, Ebel G, Grant DS, Happi CT, Park DJ, Gnirke A, Sabeti PC, Matranga CB. Capturing sequence diversity in metagenomes with comprehensive and scalable probe design. Nat Biotechnol [Internet]. 2019;37:160–168. Available from: http://www.nature.com/articles/s41587-018-0006-x

14. Wood DE, Salzberg SL. Kraken: ultrafast metagenomic sequence classification using exact alignments. Genome Biol [Internet]. 2014;15:R46. Available from: http://genomebiology.biomedcentral.com/articles/10.1186/gb-2014-15-3-r46

15. Vasimuddin M, Misra S, Li H, Aluru S. Efficient Architecture-Aware Acceleration of BWA-MEM for Multicore Systems [Internet]. In: 2019 IEEE International Parallel and Distributed Processing Symposium (IPDPS). IEEE; 2019. p. 314–324.Available from: https://ieeexplore.ieee.org/document/8820962/

16. Kim D, Paggi JM, Park C, Bennett C, Salzberg SL. Graph-based genome alignment and genotyping with HISAT2 and HISAT-genotype. Nat Biotechnol [Internet]. 2019;37:907–915. Available from: https://www.nature.com/articles/s41587-019-0201-4

17. Gel B, Serra E. karyoploteR: an R/Bioconductor package to plot customizable genomes displaying arbitrary data. Bioinformatics [Internet]. 2017;33:3088–3090. Available from: https://academic.oup.com/bioinformatics/article/33/19/3088/3857734

18. Kühl U, Lassner D, Pauschinger M, Gross UM, Seeberg B, Noutsias M, Poller W, Schultheiss H -P. Prevalence of erythrovirus genotypes in the myocardium of patients with dilated cardiomyopathy. J Med Virol [Internet]. 2008;80:1243–1251. Available from: https://onlinelibrary.wiley.com/doi/10.1002/jmv.21187

19. Schultheiss H-P, Bock C-T, Aleshcheva G, Baumeier C, Poller W, Escher F. Interferon-β Suppresses Transcriptionally Active Parvovirus B19 Infection in Viral Cardiomyopathy: A Subgroup Analysis of the BICC-Trial. Viruses [Internet]. 2022;14:444. Available from: https://www.mdpi.com/1999-4915/14/2/444

20. Schultheiss H-P, Bock T, Pietsch H, Aleshcheva G, Baumeier C, Fruhwald F, Escher F. Nucleoside Analogue Reverse Transcriptase Inhibitors Improve Clinical Outcome in Transcriptional Active Human Parvovirus B19-Positive Patients. J Clin Med [Internet]. 2021;10:1928. Available from: https://www.mdpi.com/2077-0383/10/9/1928

21. KuLJhl U, Pauschinger M, Seeberg B, Lassner D, Noutsias M, Poller W, Schultheiss H-P. Viral Persistence in the Myocardium Is Associated With Progressive Cardiac Dysfunction. Circulation. 2005;112:1965–1970.

22. KuLJhl U, Pauschinger M, Schwimmbeck PL, Seeberg B, Lober C, Noutsias M, Poller W, Schultheiss H-P. Interferon-β Treatment Eliminates Cardiotropic Viruses and Improves Left Ventricular Function in Patients With Myocardial Persistence of Viral Genomes and Left Ventricular Dysfunction. Circulation [Internet]. 2003;107:2793–2798. Available from: https://www.ahajournals.org/doi/10.1161/01.CIR.0000072766.67150.51

23. Bennett JM, Glaser R, Malarkey WB, Beversdorf DQ, Peng J, Kiecolt-Glaser JK. Inflammation and reactivation of latent herpesviruses in older adults. Brain Behav Immun [Internet]. 2012;26:739–746. Available from: https://linkinghub.elsevier.com/retrieve/pii/S0889159111005988

24. Chimenti C, Russo MA, Frustaci A. Immunosuppressive therapy in virus-negative inflammatory cardiomyopathy: 20-year follow-up of the TIMIC trial. Eur Heart J [Internet]. 2022;43:3463–3473. Available from: https://academic.oup.com/eurheartj/article/43/36/3463/6643511

25. Takeuchi S, Kawada J, Okuno Y, Horiba K, Suzuki T, Torii Y, Yasuda K, Numaguchi A, Kato T, Takahashi Y, Ito Y. Identification of potential pathogenic viruses in patients with acute myocarditis using next-generation sequencing. J Med Virol [Internet]. 2018;90:1814–1821. Available from: https://onlinelibrary.wiley.com/doi/10.1002/jmv.25263

26. Heidecker B, Williams SH, Jain K, Oleynik A, Patriki D, Kottwitz J, Berg J, Garcia JA, Baltensperger N, Lovrinovic M, Baltensweiler A, Mishra N, Briese T, Hanson PJ, Lauten A, Poller W, Leistner DM, Landmesser U, Enseleit F, McManus B, Lüscher TF, Lipkin WI. Virome Sequencing in Patients With Myocarditis. Circ Hear Fail [Internet]. 2020;13. Available from: https://www.ahajournals.org/doi/10.1161/CIRCHEARTFAILURE.120.007103

27. Nguyen QT, Sifer C, Schneider V, Allaume X, Servant A, Bernaudin F, Auguste V, Garbarg-Chenon A. Novel Human Erythrovirus Associated with Transient Aplastic Anemia. J Clin Microbiol [Internet]. 1999;37:2483–2487. Available from: https://journals.asm.org/doi/10.1128/JCM.37.8.2483-2487.1999

28. Isaacs SR, Kim KW, Cheng JX, Bull RA, Stelzer-Braid S, Luciani F, Rawlinson WD, Craig ME. Amplification and next generation sequencing of near full-length human enteroviruses for identification and characterisation from clinical samples. Sci Rep [Internet]. 2018;8:11889. Available from: https://www.nature.com/articles/s41598-018-30322-y

29. Escher F, Kühl U, Lassner D, Poller W, Westermann D, Pieske B, Tschöpe C, Schultheiss H-P. Long-term outcome of patients with virus-negative chronic myocarditis or inflammatory cardiomyopathy after immunosuppressive therapy. Clin Res Cardiol [Internet]. 2016;105:1011–1020. Available from: http://link.springer.com/10.1007/s00392-016-1011-z

30. Grzechocińska J, Tymińska A, Giordani AS, Wysińska J, Ostrowska E, Baritussio A, Caforio ALP, Grabowski M, Marcolongo R, Ozierański K. Immunosuppressive Therapy of Biopsy-Proven, Virus-Negative, Autoimmune/Immune-Mediated Myocarditis—Focus on Azathioprine: A Review of Existing Evidence and Future Perspectives. Biology (Basel) [Internet]. 2023;12:356. Available from: https://www.mdpi.com/2079-7737/12/3/356

31. McDonagh TA, Metra M, Adamo M, Gardner RS, Baumbach A, Böhm M, Burri H, Butler J, Čelutkienė J, Chioncel O, Cleland JGF, Coats AJS, Crespo-Leiro MG, Farmakis D, Gilard M, Heymans S, Hoes AW, Jaarsma T, Jankowska EA, Lainscak M, Lam CSP, Lyon AR, McMurray JJ V, Mebazaa A, Mindham R, Muneretto C, Francesco Piepoli M, Price S, Rosano GMC, Ruschitzka F, Kathrine Skibelund A, de Boer RA, Christian Schulze P, Abdelhamid M, Aboyans V, Adamopoulos S, Anker SD, Arbelo E, Asteggiano R, Bauersachs J, Bayes-Genis A, Borger MA, Budts W, Cikes M, Damman K, Delgado V, Dendale P, Dilaveris P, Drexel H, Ezekowitz J, Falk V, Fauchier L, Filippatos G, Fraser A, Frey N, Gale CP, Gustafsson F, Harris J, Iung B, Janssens S, Jessup M, Konradi A, Kotecha D, Lambrinou E, Lancellotti P, Landmesser U, Leclercq C, Lewis BS, Leyva F, Linhart A, Løchen M-L, Lund LH, Mancini D, Masip J, Milicic D, Mueller C, Nef H, Nielsen J-C, Neubeck L, Noutsias M, Petersen SE, Sonia Petronio A, Ponikowski P, Prescott E, Rakisheva A, Richter DJ, Schlyakhto E, Seferovic P, Senni M, Sitges M, Sousa-Uva M, Tocchetti CG, Touyz RM, Tschoepe C, Waltenberger J, Adamo M, Baumbach A, Böhm M, et al. 2021 ESC Guidelines for the diagnosis and treatment of acute and chronic heart failure. Eur Heart J [Internet]. 2021;42:3599–3726. Available from: https://academic.oup.com/eurheartj/article/42/36/3599/6358045

32. Schultheiss H-P, Piper C, Sowade O, Waagstein F, Kapp J-F, Wegscheider K, Groetzbach G, Pauschinger M, Escher F, Arbustini E, Siedentop H, Kuehl U. Betaferon in chronic viral cardiomyopathy (BICC) trial: Effects of interferon-β treatment in patients with chronic viral cardiomyopathy. Clin Res Cardiol [Internet]. 2016;105:763–773. Available from: http://link.springer.com/10.1007/s00392-016-0986-9

33. Kühl U, Lassner D, Wallaschek N, Gross UM, Krueger GRF, Seeberg B, Kaufer BB, Escher F, Poller W, Schultheiss H. Chromosomally integrated human herpesvirus 6 in heart failure: prevalence and treatment. Eur J Heart Fail [Internet]. 2015;17:9–19. Available from: https://onlinelibrary.wiley.com/doi/10.1002/ejhf.194

34. Tschöpe C, Ammirati E, Bozkurt B, Caforio ALP, Cooper LT, Felix SB, Hare JM, Heidecker B, Heymans S, Hübner N, Kelle S, Klingel K, Maatz H, Parwani AS, Spillmann F, Starling RC, Tsutsui H, Seferovic P, Van Linthout S. Myocarditis and inflammatory cardiomyopathy: current evidence and future directions. Nat Rev Cardiol [Internet]. 2021;18:169–193. Available from: https://www.nature.com/articles/s41569-020-00435-x

